# Physician- and Large Language Model-Generated Hospital Discharge Summaries: A Blinded, Comparative Quality and Safety Study

**DOI:** 10.1101/2024.09.29.24314562

**Authors:** Christopher Y.K. Williams, Charumathi Raghu Subramanian, Syed Salman Ali, Michael Apolinario, Elisabeth Askin, Peter Barish, Monica Cheng, W. James Deardorff, Nisha Donthi, Smitha Ganeshan, Owen Huang, Molly A. Kantor, Andrew R. Lai, Ashley Manchanda, Kendra A. Moore, Anoop N. Muniyappa, Geethu Nair, Prashant P. Patel, Lekshmi Santhosh, Susan Schneider, Shawn Torres, Michi Yukawa, Colin C. Hubbard, Benjamin I. Rosner

## Abstract

**Importance:** High quality discharge summaries are associated with improved patient outcomes but contribute to clinical documentation burden. Large language models (LLMs) provide an opportunity to support physicians by drafting discharge summary narratives.

**Objective:** To determine whether LLM-generated discharge summary narratives are of comparable quality and safety to those of physicians.

**Design:** Cross-sectional study.

**Setting:** University of California, San Francisco.

**Participants:** 100 randomly selected Inpatient Hospital Medicine encounters of 3-6 days duration between 2019-2022.

**Exposure:** Blinded evaluation of physician- and LLM-generated narratives was performed in duplicate by 22 attending physician reviewers.

**Main Outcomes and Measures:** Narratives were reviewed for overall quality, reviewer preference, comprehensiveness, concision, coherence, and three error types – inaccuracies, omissions, and hallucinations. Each error individually, and each narrative overall, were assigned potential harmfulness scores on a 0-7 adapted AHRQ scale.

**Results:** Across 100 encounters, LLM- and physician-generated narratives were comparable in overall quality on a 1-5 Likert scale (average 3.67 [SD 0.49] vs 3.77 [SD 0.57], p=0.213) and reviewer preference (χ2 = 5.2, p=0.270). LLM-generated narratives were more concise (4.01 [SD 0.37] vs. 3.70 [SD 0.59]; p<0.001) and more coherent (4.16 [SD 0.39] vs. 4.01 [SD 0.53], p=0.019) than their physician-generated counterparts, but less comprehensive (3.72 [SD 0.58] vs. 4.13 [SD 0.58]; p<0.001). LLM-generated narratives contained more unique errors (average 2.91 [SD 2.54] errors per summary) than physician-generated narratives (1.82 [SD 1.94]). Averaged across individual errors, there was no significant difference in the potential for harm between LLM- and physician-generated narratives (1.35 [SD 1.07] vs 1.34 [SD 1.05], p=0.986). Both LLM- and physician-generated narratives had low overall potential for harm (<1 on 0-7 scale), although LLM-generated narratives scored higher than physician narratives (0.84 [SD 0.98] vs 0.36 [SD 0.70], p<0.001).

**Conclusions and Relevance:** In this cross-sectional study of 100 inpatient Hospital Medicine encounters, LLM-generated discharge summary narratives were of similar quality, and were preferred equally, to those generated by physicians. LLM-generated summaries were more likely to contain errors but had low overall harmfulness scores. Our findings suggest that LLMs could be used to draft discharge summary narratives of comparable quality and safety to those written by physicians.

**Key Points:** *Question:* Can large language models (LLMs) draft hospital discharge summary narratives of comparable quality and safety to those written by physicians?

*Findings:* In this cross-sectional study of 100 discharge summaries, LLM- and physician-generated narratives were rated comparably by blinded reviewers on overall quality and preference. LLM-generated narratives were more concise and coherent than their physician-generated counterparts, but less comprehensive. While LLM-generated narratives were more likely to contain errors, their overall potential for harm was low.

*Meaning:* These findings suggest the potential for LLMs to aid clinicians by drafting discharge summary narratives.

## Introduction

The hospital discharge summary is a form of clinical documentation essential for facilitating a patient’s safe transition from the hospital to the post-acute setting.^1^ High-quality discharge summaries are associated with reduced medication errors, lower hospital readmission rates, and enhanced primary care physician (PCP) satisfaction.^2–7^ The Transitions of Care Consensus Policy Statement recommends that high-quality discharge summaries contain elements including principal diagnosis and problem list, medication list, test results, and others.^8^ With the advent of the electronic health record, templated discharge summaries have facilitated the automated completion of several of these components.

However, composing the discharge summary narrative sections, including the history of the presenting illness and the hospital course, remains a time-consuming process, a substantial contributor to documentation burden, and a potential detractor from face-to-face patient care.^9,10^ Unlike a hospital progress note which often reflects incremental daily documentation effort, a discharge summary can be considerably more involved, particularly for lengthy hospital encounters or when care has been provided by sequential physicians, the last of whom must reconstruct the salient encounter events. In one report, 44% of hospitalists described being too busy to prepare high-quality discharge summaries.^11^ From the perspective of the discharge summary recipient – often the PCP or a skilled nursing facility (SNF) physician – content deficits are common.^3,11^ Furthermore, differences of opinion between hospital physicians and PCPs exist as to what constitutes an appropriately comprehensive narrative.^11^

The emergence of large language models (LLMs) such as Generative Pre-Trained Transformer (GPT), a form of artificial intelligence (AI) capable of reviewing large quantities of information and synthesizing original content emulating human composition, offers promise in healthcare.^12–19^ Given increasing physician documentation burden, there is now opportunity to use LLMs to draft narrative components of the discharge summary for the physician to review, akin to the manner in which LLMs are being used to draft clinical notes and inbox message responses to patients.^13,20^ Existing studies of LLM clinical text summarization – the task of producing a shorter version of a clinical document, while preserving information content and remaining faithful to the source – have largely focused on text from Emergency Department notes, radiology reports, and doctor-patient conversations.^20–25^ However, the summarization of multiple documents from real-world inpatient encounters written by different healthcare providers, as is required to generate the hospital discharge narrative, is a more complex task. While neither LLMs nor the healthcare system may be ready to fully replace the clinician’s involvement with the discharge narrative, LLMs may offer opportunities to reduce clinician burden by drafting narratives to be reviewed and edited. Therefore, evaluating LLM performance on this task for quality and safety is essential before clinical implementation.

In this study, we sought to investigate the quality and safety of discharge summary narratives for real-world, inpatient Hospital Medicine encounters generated by an LLM from the corpus of all hospital encounter notes. Using standardized quality and safety metrics, we compared LLM- to physician-generated narratives for the same encounter, incorporating the blinded reviews of both discharge summary producers (hospitalists) and common consumers (PCPs and SNF physicians).

## Methods

The University of California San Francisco (UCSF) Information Commons contains structured clinical data and text notes for over 950,000 inpatient encounters from 2012 to 2024.^26^ The UCSF Institutional Review Board determined that use of these deidentified data is exempt from approval and informed consent. This study was conducted according to a pre-specified protocol (Supplementary File 1).

### Study Cohort

We identified historical hospital encounters and their corresponding clinical notes for patients who received care under the UCSF Hospital Medicine service between 2019-2022. We limited encounter durations to 3-6 days as both a proof of concept and to mitigate the burden on reviewers for reading the entire set of inpatient notes for longer encounters. Additional inclusion criteria consisted of encounters exclusively under the Hospital Medicine service (i.e. no transfers between specialties) and patients who were discharged alive. Exclusion criteria consisted of encounters lacking clinical notes (e.g. administrative encounters) or lacking available discharge summaries, and encounters with discharge summaries written by non-physicians (rare in Hospital Medicine at UCSF). Due to GPT-4 context window limits, encounters in which the corpus of encounter note text exceeded 31,000 tokens were also excluded (Figure 1). During initial study design and cohort selection, the GPT-4 model available had a 32,000 token limit. GPT-4 Turbo, with a 128,000 token limit, became available after finalizing cohort selection and was used for summarization. Where more than one discharge summary was available, the latest was selected.

**Figure 1.**
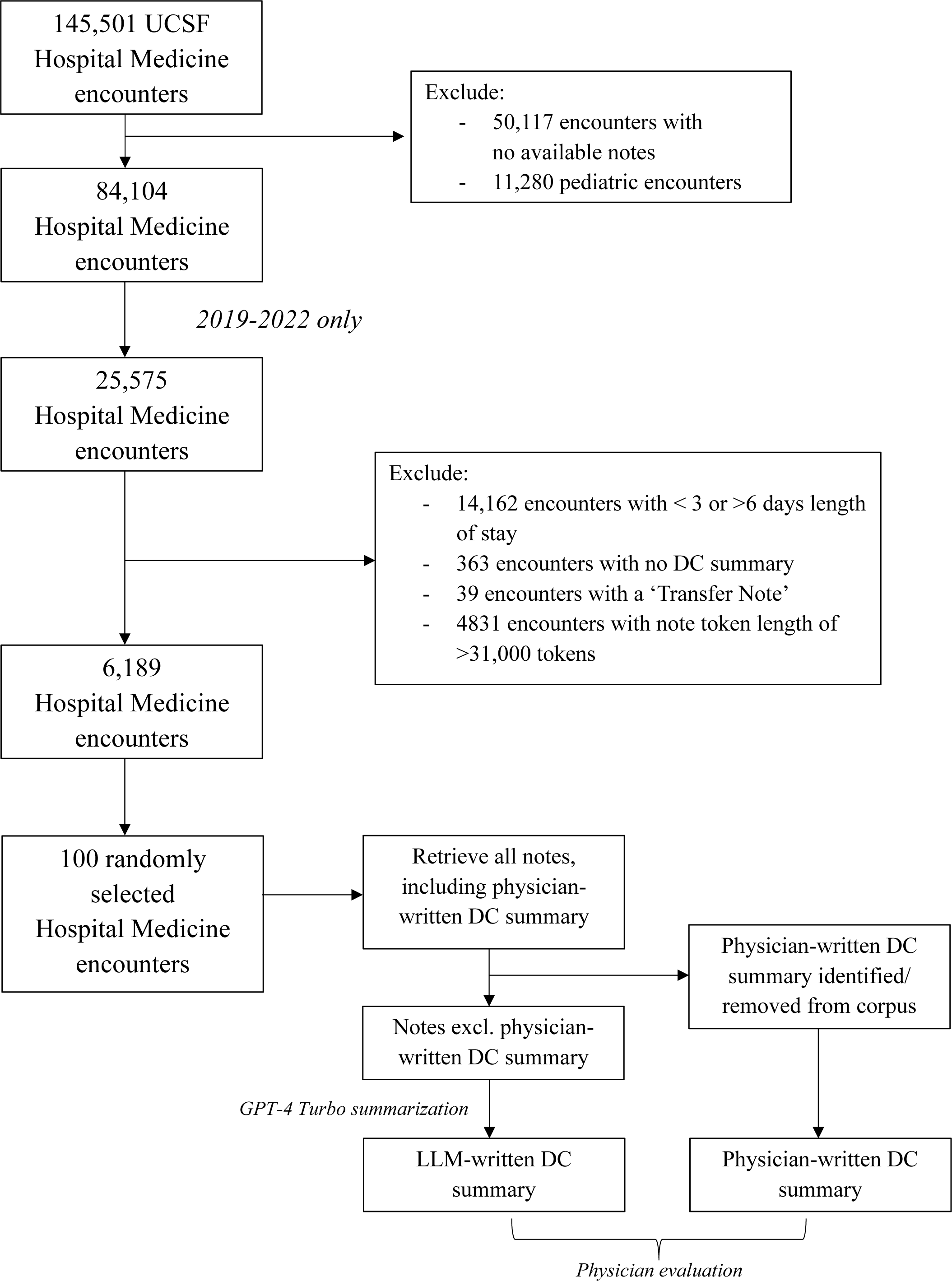
Flowchart of included Hospital Medicine encounters. DC = Discharge.

### Encounter Note Processing

Software was written using regular expressions to examine the section headings of discharge summaries from the cohort encounters. The narrative section was extracted, corresponding to the text from the ‘Admission Diagnosis’ heading to the ‘Physical Exam’ heading (Supplementary File 2). We next retrieved all the preceding encounter clinical notes unrelated to the patient’s discharge. These notes served as both the corpus of input text for LLM summarization and the reference text against which both LLM- and physician-generated narratives were evaluated by reviewers (Figure 1).

### Generation of Discharge Summary Narratives

From the dataset of cohort encounters meeting inclusion and exclusion criteria, we randomly sampled an n = 100 test set for evaluation, alongside a separate n = 100 development set for prompt engineering and reviewer training (Supplementary Files 1 and 3). Using UCSF’s secure, HIPAA-compliant Versa Application Programming Interface (API) on Microsoft Azure, we prompted GPT-4 Turbo (model *GPT-4-turbo-128K*, temperature = 0, other settings as default) to summarize the concatenated corpus of clinical notes into a discharge summary narrative for each patient’s encounter.

### Discharge Summary Narrative Evaluation

#### Qualitative Evaluation

Evaluation of both physician- and LLM-generated narratives was performed using a two-part approach that included reviews by 14 attending Hospital Medicine physicians (hospitalists), 3 PCPs, and 5 SNF physicians (non-hospitalists) (Supplementary Table 1). Reviewers were blinded to which narrative was physician-vs. LLM-generated. All metrics, and the reviewer types responsible for each metric, are displayed in Supplementary Table 2 and Supplementary Figure 1.

First, the 14 hospitalists reviewed both physician- and LLM-generated narratives for errors. Two hospitalists were randomly assigned to separately review these narratives for each of 14-15 study cohort encounters. Rather than assessing the LLM-generated narrative against the physician-generated narrative as a reference, we used the full corpus of encounter notes as the reference for both, enabling comparison of error rates between LLM-*and* physician-generated narratives. Reviewers were instructed to classify errors into three types commonly described in the literature - inaccuracies, omissions, and hallucinations (see Supplementary File 1 for definitions).^22,24,27^ Reviewers then rated the potential for harm from each error using the Agency for Healthcare Research and Quality (AHRQ) Common Format Harm Scale adapted to reflect the *potential* for harm rather than *actual* harm (Supplementary File 1).^28^ Reviewers additionally gave each narrative an overall potential harmfulness score.

Prior to reviewing the 100 study cohort encounters, reviewers underwent training and evaluation on error classification using two development set encounters. Eight reviewers met retraining criteria as described in the study protocol (Supplementary Files 1 and 3). After training, all 100 study cohort encounters were reviewed for errors by two independent reviewers (a common approach in patient safety research),^29–31^ resulting in an initial 200 reviews. To create a list of unique errors for each narrative, a third adjudicator (BR) then merged duplicate errors and averaged harmfulness scores across duplicates.

All reviewers (hospitalists and non-hospitalists) evaluated both physician- and LLM-generated narratives on 5-point Likert scales for overall quality, and on the following three global metrics: comprehensiveness, concision, and coherence (see Supplementary File 1 for definitions and details).^24,32,33^ Reviewers also indicated a preference between the two narratives (one over the other or both considered ‘equal preference’). Hence, global scores for each encounter were assessed by four reviewers in total: two hospitalists (as part of their reviews of errors above) and two randomly assigned non-hospitalists.

#### Quantitative Evaluation

Because human evaluation of LLM output does not readily scale, we sought to additionally characterize the likeness of LLM- and physician-generated narratives using BLEU, ROUGE-L, METEOR, and cosine similarity scores.^34–36^ The latter were derived by calculating the cosine similarity between embeddings for the LLM- and corresponding physician-generated narrative, using the text-embedding-ada-002 model.^37^ For each encounter, we also examined the association between these quantitative metrics and the global scores assigned by reviewers.

While these metrics are widely used in the natural language processing community, their utility applied to clinical text remains unclear.^23,24^ To better understand the baseline values of these metrics for narratives from *unrelated* encounters, as well as the extent to which each metric evaluated the narrative content rather than its structure or general terminology, we additionally calculated each metric comparing the LLM-generated narrative with that of a randomly selected alternative physician-generated narrative from the cohort.

### Statistical Analysis

Mean unique error counts per narrative (overall and stratified by error type) and global rating scores (overall and stratified by reviewer type: hospitalist, PCP, SNF physician) were compared using the Wilcoxon signed-rank test against the null hypothesis of no significant difference between physician- and LLM-generated narratives. Individual error harmfulness scores were compared using the Mann-Whitney U test, as were BLEU, ROUGE-L, METEOR and cosine similarity scores. Categorical variables were compared using the chi-square test. P <0.05 was considered significant. Analyses were performed in Python and R.

## Results

### Study Cohort

Of the 145,501 Hospital Medicine encounters in the clinical data warehouse, 6,189 met inclusion criteria (Figure 1). A random n=100 sample was selected for GPT-4 Turbo summarization and reviewer evaluation. Demographic and clinical characteristics of patients associated with these encounters are displayed in Supplementary Tables 3 and 4.

### Discharge Summary Narrative Evaluation

#### Qualitative Evaluation: Individual Errors and Harm

Across 100 encounters, there were an average of 2.91 (SD 2.54) unique errors per LLM-generated narrative and 1.82 (SD 1.94) per physician-generated narrative (p<0.001). LLM-generated narratives had a greater average number of inaccuracies (0.93 [SD 0.99] vs 0.65 [SD 1.01], p=0.013) and omissions (1.75 [SD 2.09] vs 0.86 [SD 1.43], p<0.001) than physician-generated narratives (Figure 2). In contrast, LLM-and physician-generated narratives contained a similar number of hallucination errors (0.23 [SD 0.51] vs. 0.31 [SD 0.53], p=0.230).

**Figure 2.**
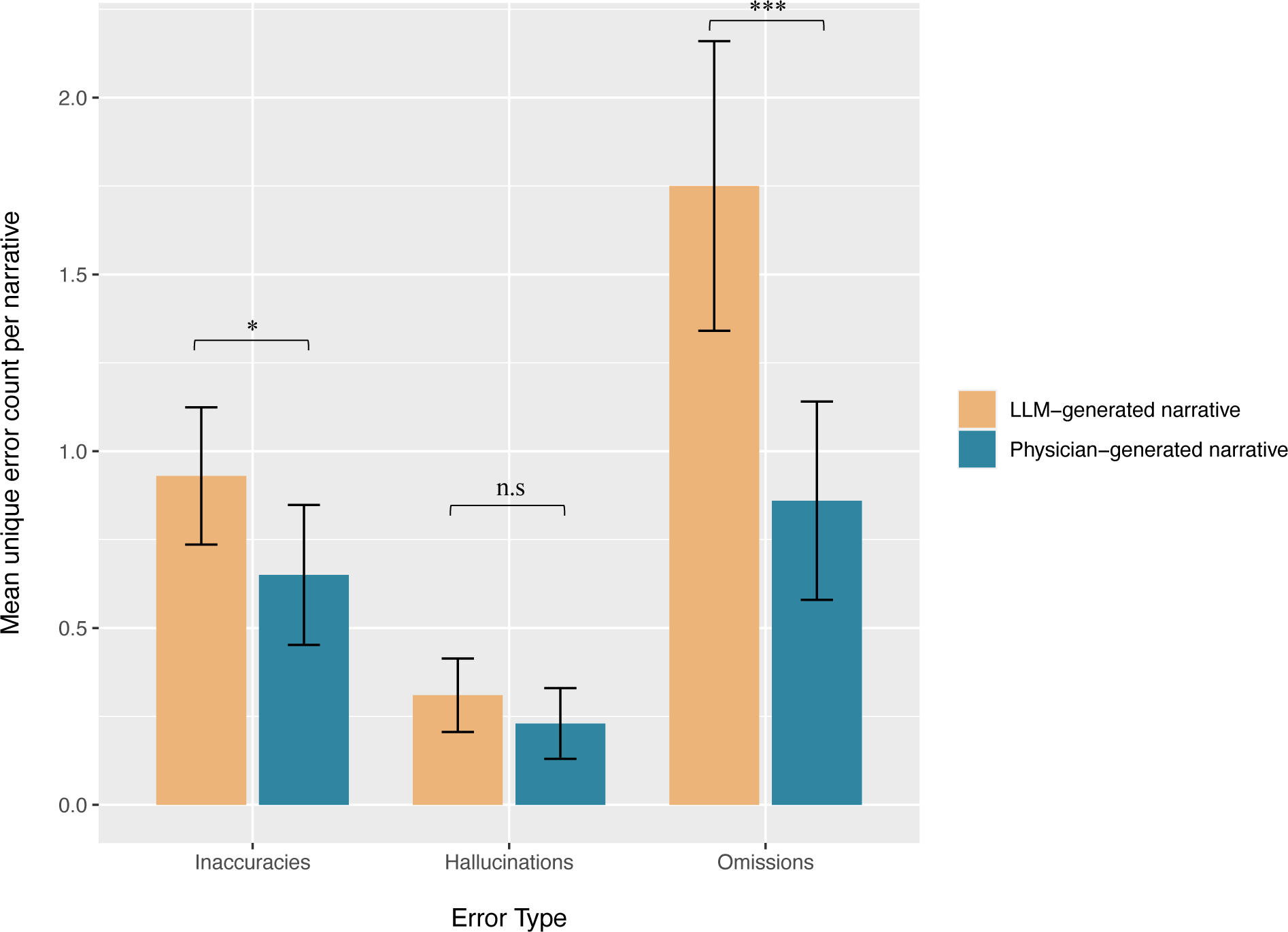
Mean unique error counts per narrative and 95% confidence intervals for each error type, averaged across 100 LLM-generated and physician-generated discharge summary narratives. n.s = Not significant; * < 0.05, ** < 0.01, *** < 0.001 (Wilcoxon signed-rank test).

Across all error types, there was no significant difference in the potential for harm per error between LLM- and physician-generated narratives (mean harmfulness score 1.35 [SD 1.07] vs. 1.34 [SD 1.05]; p=0.986). Similarly, there were no significant differences in harmfulness scores when errors were stratified by error type (Table 1; Supplementary Figure 2). Among LLM-generated narratives, there were six errors assigned potential harmfulness scores of 4 (‘Potential for permanent harm’) or greater, including five omissions and one inaccuracy (Supplementary Table 5). Physician-generated narratives contained 5 errors with potential harmfulness scores of 4 (all omissions).

**Table 1.**
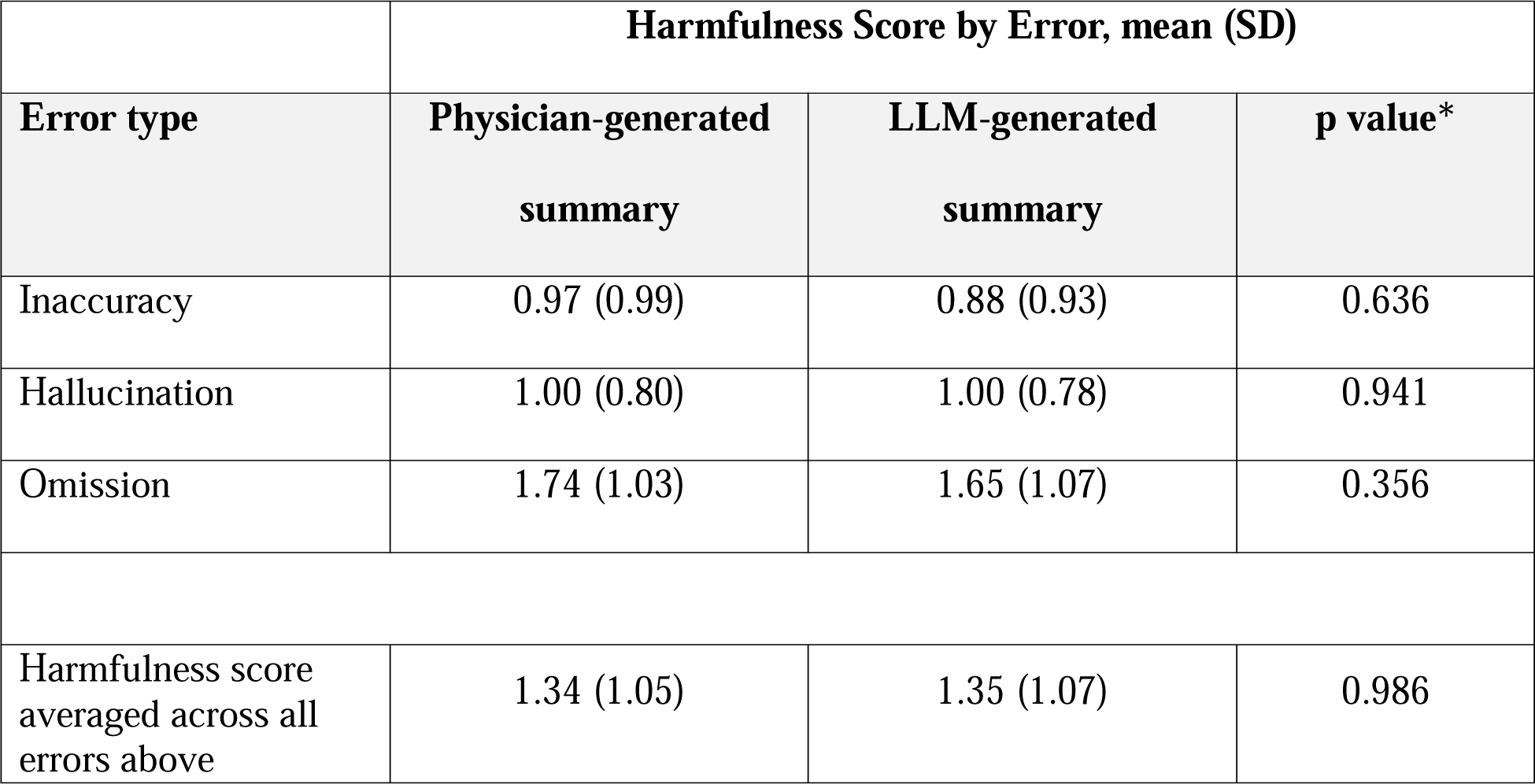
Mean harmfulness scores of individual errors identified in physician- and LLM-generated discharge summary narratives based on adapted AHRQ Common Format Harm Scale: 0 – No potential for harm, 1 – Potential for emotional distress or inconvenience (mild and transient anxiety or pain or physical discomfort), 2 – potential for requiring additional treatment, 3 – Potential for temporary harm (bodily or psychological injury, but likely not permanent), 4 – Potential for permanent harm (lifelong bodily or psychological injury or increased susceptibility to disease), 5 – Potential for lifelong bodily or psychological injury or disfigurement, 6 – Potential for severe permanent harm, 7 – potential for death. *Mann-Whitney U test; p<0.05 considered significant.

|  | Harmfulness Score by Error, mean (SD) |  |  |
| --- | --- | --- | --- |
| Error type | Physician-generated<br>summary | LLM-generated<br>summary | p value* |
| Inaccuracy | 0.97 (0.99) | 0.88 (0.93) | 0.636 |
| Hallucination | 1.00 (0.80) | 1.00 (0.78) | 0.941 |
| Omission | 1.74 (1.03) | 1.65 (1.07) | 0.356 |
| Harmfulness score averaged across all errors above | 1.34 (1.05) | 1.35 (1.07) | 0.986 |

#### Qualitative Evaluation: Global Metrics

In aggregate, hospitalists, PCPs, and SNF physicians rated GPT-generated narratives as more concise (4.01 [SD 0.37] vs. 3.70 [SD 0.59]; p<0.001), more coherent (4.16 [SD 0.39] vs. 4.01 [SD 0.53], p=0.019), but less comprehensive (3.72 [SD 0.58] vs. 4.13 [SD 0.58]; p<0.001) than their physician-generated counterparts (Table 2). As subgroups, each reviewer type found LLM-generated narratives less comprehensive than physician-generated narratives. Both PCP (3.51 [SD 0.96] vs. 3.07 [SD 1.13], p = 0.021) and SNF physicians (4.18 [SD 0.71] vs. 3.62 [SD 0.96], p<0.001) rated LLM-generated narratives as more concise, while only SNF physicians rated LLM-generated narratives more coherent (4.15 [SD 0.51] vs. 3.89 [SD 0.70]; p = 0.005). Unlike the harmfulness scores associated with individual errors (no difference), LLM-generated narratives were viewed as more harmful overall than their physician-generated counterparts (mean global harmfulness score 0.84 [SD 0.98] vs. 0.36 [SD 0.70] on a 0-7 scale; p<0.001), although both narratives’ average scores were below ‘1: Potential for emotional distress or inconvenience (mild and transient anxiety or pain or physical discomfort)’ (Table 2; Supplementary Figure 3).

**Table 2.** Mean (SD) comprehensiveness, concision, coherence, overall harmfulness, and global rating scores for physician- and LLM-generated discharge summary narratives, stratified by reviewer type. PCP = primary care physician, SNF = skilled nursing facility physician. N/a for non-hospitalist reviewers as only hospitalists reviewed errors against the reference corpus of hospital encounter notes. *Wilcoxon signed-rank test; p<0.05 considered significant. **5-point Likert scale: 1 - Strongly disagree, 2 - Disagree, 3 - Neutral, 4 - Agree, and 5 - Strongly agree. ***Adapted AHRQ Common Format Harm Scale consisting of options: 0 – No potential for harm, 1 – Potential for emotional distress or inconvenience (mild and transient anxiety or pain or physical discomfort), 2 – Potential for requiring additional treatment, 3 – Potential for temporary harm (bodily or psychological injury, but likely not permanent), 4 – Potential for permanent harm (lifelong bodily or psychological injury or increased susceptibility to disease), 5 – Potential for lifelong bodily or psychological injury or disfigurement, 6 – Potential for severe permanent harm, 7 – Potential for death.

| Score, mean<br>(SD) | Hospitalist, n=14 |  |  | PCP, n=3 |  |  | SNF, n=5 |  |  | All reviewers, n=22 |  |  |
| --- | --- | --- | --- | --- | --- | --- | --- | --- | --- | --- | --- | --- |
|  | *Physician* | *LLM* | *p*<br>*value\** | *Physician* | *LLM* | *p*<br>*value\** | *Physician* | *LLM* | *p*<br>*value\** | *Physician* | *LLM* | *p*<br>*value\** |
| Comprehensiveness** | 4.20<br>(0.72) | 3.73<br>(0.8) | <0.001 | 3.93<br>(1.19) | 3.32<br>(1.32) | 0.004 | 4.18<br>(0.75) | 3.95<br>(0.67) | 0.015 | 4.13<br>(0.58) | 3.72<br>(0.58) | <0.001 |
| Concision** | 3.99<br>(0.73) | 4.12<br>(0.60) | 0.246 | 3.07<br>(1.13) | 3.51<br>(0.96) | 0.021 | 3.62<br>(0.96) | 4.18<br>(0.71) | <0.001 | 3.70<br>(0.59) | 4.01<br>(0.37) | <0.001 |
| Coherence** | 4.29<br>(0.64) | 4.35<br>(0.56) | 0.495 | 3.53<br>(1.26) | 3.75<br>(1.15) | 0.313 | 3.89<br>(0.70) | 4.15<br>(0.51) | <0.001 | 4.01<br>(0.53) | 4.16<br>(0.39) | 0.019 |
| Global harmfulness*** | 0.36<br>(0.70) | 0.84<br>(0.98) | <0.001 | *n/a* | *n/a* | *n/a* | *n/a* | *n/a* | *n/a* | *n/a* | *n/a* | *n/a* |
| Global quality rating** | 4.04<br>(0.69) | 3.76<br>(0.71) | 0.007 | 3.29<br>(1.19) | 3.17<br>(1.21) | 0.591 | 3.67<br>(0.87) | 3.89<br>(0.70) | 0.054 | 3.77<br>(0.57) | 3.67<br>(0.49) | 0.213 |

Overall, there was no significant difference between LLM- and physician-generated narratives in the mean overall quality rating (3.67 [SD 0.49] vs 3.77 [SD 0.57]; p=0.674) (Table 2) or in reviewer preference (Table 3; χ2 = 5.2, p=0.27).

**Table 3.** Preference counts detailing which of the physician- or LLM-generated discharge summary narratives reviewers preferred overall, stratified by reviewer type. PCP = primary care physician, SNF = skilled nursing facility physician. χ^2^ = 5.2, p=0.270.

| Preference | Count (%) |  |  | Total (%) |
| --- | --- | --- | --- | --- |
|  | *Hospitalist* | *PCP* | *SNF* |  |
| Physician-generated summary | 104 (52%) | 39 (52%) | 52 (42%) | 195 (49%) |
| LLM-generated summary | 64 (32%) | 28 (37%) | 51 (41%) | 143 (36%) |
| Equal preference | 32 (16%) | 8 (11%) | 22 (18%) | 62 (16%) |

#### Quantitative Metrics

There was low correlation between quantitative metrics and reviewer global scores (Supplementary Table 6). While similarity metrics between LLM- and physician-generated narratives were significantly higher for the same encounter than for different encounters (Supplementary Table 7), the absolute values of the ROUGE-L, BLEU, and METEOR scores, even for the same encounters, were low. On the other hand, although there was a high average cosine similarity between physician- and LLM-generated narratives for the same encounter, its baseline (a randomly selected, alternative encounter) was also high, suggesting that cosine similarity may be of limited evaluative utility for clinical text summarization.

## Discussion

We conducted a blinded cross-sectional study of 100 physician-versus LLM-generated discharge summary narratives for quality and safety as an appropriate first step towards assessing the role of LLMs in drafting these narratives in real-world practice. Overall, we found no differences in either quality or reviewer preference between physician- and LLM-generated narratives.

Our results suggest that neither physicians nor LLMs consistently write ‘perfect’ narratives. Although LLM-generated narratives were more likely to contain errors (particularly greater omissions and inaccuracies), physician narratives were just as likely to contain hallucination type errors. This is notable given the well-documented propensity for LLMs to hallucinate.^38,39^ One possible contributing factor to this finding may be the availability of new information to the physician on writing the discharge summary, which was not previously documented in the notes. However, we cannot discount that human fallibility in reconstructing historical events over the course of the encounter could play a role.^23^

There was no difference in potential for harm at the individual error level between LLMs and physicians. However, reviewers found that the overall potential for harm was greater in LLM-generated narratives, perhaps because of the cumulative effect of more errors in LLM-generated narratives. Nevertheless, in both cases (average scores of 0.36 and 0.84 out of 7 in physician and LLM narratives, respectively) this potential for harm was extremely low (less than ‘Potential for emotional distress or inconvenience’ on the adapted AHRQ Common Format Harm Scale). Differences in perception of the meaning of the harm scale units among healthcare providers suggest that this half-point difference may have limited clinical significance.^28,40^ Only one LLM-generated narrative scored 4 (‘Potential for permanent harm’) or higher, suggesting a low overall potential for harm across both physician- and LLM-generated narratives.

LLM-generated narratives were more coherent and concise, but less comprehensive than their physician-generated counterparts. The respective scores for comprehensiveness and concision across LLM and physician narratives, paired with the differences in omission rates of each, are unsurprising. This likely reflects opposing sides of the summarization spectrum, whereby a more concise summary is more likely to omit key details and consequently lack comprehensiveness. It is possible that, with more focused guidance, LLMs can be prompted to make fewer errors of omission and therefore increase the comprehensiveness of their narratives, albeit at the risk of compromising concision.

Our findings highlight the potential use of LLMs to draft hospital discharge summary narratives for clinician review and editing that are of comparable quality and safety to physician-generated narratives. This is an important first step as LLMs begin to be deployed in clinical practice.^20^ Previous research on LLM clinical text summarization has demonstrated promising results when the summarization task is for single documents.^22–24,41^ For example, one study of four clinical summarization tasks – radiology reports, patient questions, progress notes, and doctor-patient dialogue – found that LLM-generated summaries were equivalent or superior to those generated by medical experts.^23^ While evaluating LLMs for discharge summarization is relatively nascent, several studies have been reported.^22,41–43^ One, using fine-tuned BERT and BART models to generate hospital discharge summaries for neurology patients, found that only 62% met the standard of care and underperformed human physicians on metrics of quality, readability, factuality, and completeness.^43^ In a study of GPT-4 Turbo’s ability to generate 53 discharge summaries using the MIMIC-III ICU dataset, although relatively high accuracy was found, over one third of errors were classified as severe omissions and nearly 15% classified as hallucinations.^42^ However, no comparison between physician- and LLM-generated summaries against the reference corpus was reported; an important approach to understanding the quality and safety of these models before real-world deployment.

The extent to which LLM-drafted discharge summary narratives may reduce clinical documentation burden and improve clinician efficiency remains unclear. Recent studies of LLM-drafted replies to patient messages demonstrate reduced clinician burnout, but no reductions in time spent reading/writing messages.^13,44^ Although our results suggest that LLM- and physician-generated narratives are of comparable quality and safety, evaluations of the impact on documentation burden and efficiency are still needed. This is particularly important given the lack of reliable automated means to identify errors in LLM-drafted narratives.^23,27,43^ Among the few errors in our study with potential harmfulness scores of 4 or greater, most were omissions, reflecting the ongoing need for physician review to ensure that all pertinent information is included in the discharge summary narrative. Ultimately, to optimize for safety and quality, while benefitting from the physician’s comprehensiveness and LLM’s concision, a clinician-in-the-loop approach to review and edit LLM-drafted narratives is likely to remain essential.

## Limitations

There are several limitations to this study. First, although the study was blinded and LLM-generated narratives were structured in a similar style to physician-generated counterparts, reviewers may have been able to surmise which narrative was which due to syntax such as the increased use of abbreviations and higher prevalence of redacted identifiable information in the physician-generated narratives. Second, our cohort included only patient encounters with a length of stay between 3-6 days, inclusive. This was a practical decision based on the LLM token limit and to reduce the review burden of full encounter content on reviewers. Consequently, the ability of LLMs to generate summary narratives for more complex encounters exceeding 6 days in length is unclear. Third, it is possible that the quality of the LLM-generated narratives could have been improved with further prompt engineering, such as more detailed guidance to prioritize summary comprehensiveness over concision discussed previously.

## Conclusions

In this cross-sectional study of 100 inpatient Hospital Medicine encounters, there were no differences in the overall quality rating or reviewer preferences between LLM- and physician-generated narratives. LLM-generated narratives were more concise and more coherent than their physician-generated counterparts, but less comprehensive. LLM-generated narratives were more likely to contain errors but had low overall potential for harm. Our findings suggest that LLMs could be used to draft discharge summary narratives of comparable quality and safety to physicians for inpatient hospital encounters.

## Data Availability

Data is not available.

## Acknowledgments

The authors acknowledge the use of the UCSF Information Commons computational research platform, developed and supported by UCSF Bakar Computational Health Sciences Institute. The authors also thank the UCSF AI Tiger Team, Academic Research Services, Research Information Technology, and the Chancellor’s Task Force for Generative AI for their software development, analytical and technical support related to the use of Versa API gateway (the UCSF secure implementation of large language models and generative AI via API gateway), Versa chat (the chat user interface), and related data asset and services. We also thank Julia Adler-Milstein, PhD for thoughtful feedback on the manuscript.

## Author Contributions

CYKW had full access to all of the data in the study and takes responsibility for the integrity of the data and the accuracy of the data analysis.

Concept and design: CYKW, CRS, BIR

Acquisition, analysis, or interpretation of data: CYKW, CRS, BIR

Drafting of the manuscript: CYKW, CRS, BIR

Critical revision of the manuscript for important intellectual content: CRS, SSA, MA, PB, ND, SG, OH, MK, AL, AM, GN, PP, LS, BIR, EA, ST, KM, MC, JD, AM, SS, MY

Statistical analysis: CYKW, CCH, BIR

Supervision: BIR

