## Supplementary File 2 for "Physician- and Large Language Model-Generated Hospital Discharge Summaries: A Blinded, Comparative Quality and Safety Study"

**MEDICAL CENTER - DISCHARGE SUMMARY**

Patient Name: Test Admit, Patient

Patient MRN: 12345

Date of Birth: 1/1/1980

Facility: ***

Attending Physician: Test, Attending Physician

Date of Admission: 3/28/2023

Date of Discharge: 4/3/2023

Admission Diagnosis: GI bleed [K92.2]

Discharge Diagnosis: Duodenal ulcer

Discharge Disposition: Home with Services

Discharge summary narrative

**History (with Chief Complaint)**

***

**Brief Hospital Course by Problem**

***

**Physical Exam at Discharge**

Temp 37.0^o^C (98.6^o^F) | Ht 178 cm (5' 10.08") | Wt 76 kg (167 lb 8.8 oz) | BMI 23.99 kg/m^2^

Net IO Since Admission: 779 mL [08/14/24 2330]

Physical Exam:

***

**Relevant Labs, Radiology, and Other Studies**

Labs: ***

No visits with results within 1 Month(s) from this visit.

Latest known visit with results is:

Imaging:

***

Micro:

***

**Procedures Performed and Complications**

***

**DISCHARGE INSTRUCTIONS**

**Discharge Diet**

***

**Functional Assessment at Discharge/Activity Goals**

***

**Allergies and Medications at Discharge**

Meds:

***

Allergies:

***

**Pending Tests**

***

**Outside Follow-up**

***

**Booked Follow-up Appointments**

No future appointments.

**Pending Referrals**

***

**Case Management Services Arranged**

**Case Management Services Arranged: (all recorded)**

**Discharge Assessment**

Condition at discharge: {Desc; good/fair/poor: }

**Covid-19 Vaccines**

|  | No immunizations on file. |
| --- | --- |

**Primary Care Physician**

Test PCP

Address: Main Street

Phone: XXX-XXX-XXXX

Fax: YYY-YYY-YYYY

Outside Providers, for pending tests please use the following numbers:

For Laboratory - Please Call: XXX-XXX-XXXX

For Microbiology - Please Call: XXX-XXX-XXXX

For Pathology - Please Call: XXX-XXX-XXXX

**Signed,**

Attending Physician Test, MD

1/1/2024

‌

‌**Discharge Instructions provided to the patient (if any):**

‌

**Patient Instructions**

***
