## Supplementary File 3 for "Physician- and Large Language Model-Generated Hospital Discharge Summaries: A Blinded, Comparative Quality and Safety Study"

**Supplementary Tables**

| **Reviewer Role** | **Author Initials** |
| --- | --- |
| Hospitalist | CRS, SSA, MA, PB, ND, SG, OH, MK, AL, AM, GN, PP, LS, BIR |
| PCP | EA, ST, KM |
| SNF physician | MC, WJD, AM, SS, MY |

**Supplementary Table 1.** Reviewer roles.

| **Metric** | **Scale** | **Reviewer Type** |
| --- | --- | --- |
| Errors against full hospital encounter corpus | | |
| Inaccuracy | Count within each narrative | Hospitalist* |
| Omission | Count within each narrative | Hospitalist* |
| Hallucination | Count within each narrative | Hospitalist* |
| Potential harmfulness of individual errors | | |
| AHRQ Common Format Harm Scale (adapted from actual harm to potential for harm) | Adapted AHRQ Common Format Harm Scale** | Hospitalist* |
| Global metrics at the narrative level | | |
| Comprehensiveness: ‘This summary is appropriately comprehensive’ | 5-point Likert scale*** | Hospitalist, PCP, SNF |
| Concision: ‘This summary is appropriately concise’ | 5-point Likert scale*** | Hospitalist, PCP, SNF |
| Coherence: ‘This summary is appropriately coherent’ | 5-point Likert scale*** | Hospitalist, PCP, SNF |
| Overall quality: ‘Overall, the narrative for this discharge summary is high quality’ | 5-point Likert scale*** | Hospitalist, PCP, SNF |
| Overall potential harmfulness | Adapted AHRQ Common Format Harm Scale** | Hospitalist* |
| Preference between LLM- or physician-generated narrative | Prefer LLM narrative *or*  Prefer physician narrative *or*  Equal preference | Hospitalist, PCP, SNF |

**Supplementary Table 2.** Evaluation metrics. *Only hospitalists evaluated each narrative for errors and potential harmfulness from errors against the full encounter notes, as they are the only ones in routine care to access the full corpus of hospital encounter notes. **Adapted AHRQ Common Format Harm Scale consisting of options: 0 – No potential for harm, 1 – Potential for emotional distress or inconvenience (mild and transient anxiety or pain or physical discomfort), 2 – Potential for requiring additional treatment, 3 – Potential for temporary harm (bodily or psychological injury, but likely not permanent), 4 – Potential for permanent harm (lifelong bodily or psychological injury or increased susceptibility to disease), 5 – Potential for lifelong bodily or psychological injury or disfigurement, 6 – Potential for severe permanent harm, 7 – Potential for death. ***1 - Strongly disagree, 2 - Disagree, 3 - Neutral, 4 - Agree, and 5 - Strongly agree. PCP = Primary Care Physician; SNF = Skilled Nursing Facility physician.

|  | **No.** |
| --- | --- |
| **Sex** | |
| Male | 47 |
| Female | 53 |
| **Race/Ethnicity** | |
| White | 48 |
| Asian | 25 |
| Black or African American | 9 |
| Latinx | 8 |
| Multi-Race/Ethnicity | 4 |
| Southwest Asian and North African | 2 |
| Other | 2 |
| Unknown/Declined | 2 |
| **Age** | |
| Median (IQR) | |
| 66.1 years (44.8 – 80.2 years) | |
| 18 (inclusive) to 30 years | 12 |
| 30 (inclusive) to 45 years | 14 |
| 45 (inclusive) to 60 years | 11 |
| 60 (inclusive) to 75 years | 25 |
| 75+ years | 38 |

**Supplementary Table 3.** Baseline demographic characteristics of sampled patient encounters.

| **Main Diagnosis-Related Group (DRG) associated with admission** | **Count** |
| --- | --- |
| SEPTICEMIA OR SEVERE SEPSIS WITHOUT MV >96 HOURS WITH MCC | 7 |
| PULMONARY EDEMA AND RESPIRATORY FAILURE | 5 |
| SEPTICEMIA AND DISSEMINATED INFECTIONS | 4 |
| SEPTICEMIA OR SEVERE SEPSIS WITHOUT MV >96 HOURS WITHOUT MCC | 4 |
| NERVOUS SYSTEM NEOPLASMS WITH MCC | 3 |
| CHEMOTHERAPY WITHOUT ACUTE LEUKEMIA AS SECONDARY DIAGNOSIS WITH CC | 3 |
| OTHER KIDNEY AND URINARY TRACT DIAGNOSES WITH MCC | 3 |
| ALCOHOL, DRUG ABUSE OR DEPENDENCE WITHOUT REHABILITATION THERAPY WITH MCC | 3 |
| MISCELLANEOUS DISORDERS OF NUTRITION, METABOLISM, FLUIDS AND ELECTROLYTES WITHOUT MCC | 3 |
| RENAL FAILURE WITH CC | 2 |
| GASTROINTESTINAL HEMORRHAGE WITH CC | 2 |
| ORGANIC DISTURBANCES AND INTELLECTUAL DISABILITY | 2 |
| RESPIRATORY INFECTIONS AND INFLAMMATIONS WITH MCC | 2 |
| PULMONARY EMBOLISM WITH MCC OR ACUTE COR PULMONALE | 2 |
| CELLULITIS WITHOUT MCC | 2 |
| HIV WITH MAJOR RELATED CONDITION WITH CC | 2 |

**Supplementary Table 4.** Clinical characteristics of (n = 100) sampled patients: top 16 most common Diagnosis-Related Group (DRG) associated with admission.

| **Summary type** | **Encounter ID** | **Error type** | **Error potential for harm score** | **Reviewer-provided explanation of score** |
| --- | --- | --- | --- | --- |
| LLM-generated | D1DF9EF3957287 | Omission | 4 | CT showed aortic ectasia which is not mentioned in the DC summary (also not mentioned in progress notes). |
| LLM -generated | D46E652402BA78 | Omission | 4 | Neurosurgery recommended C collar for 6 weeks post op but DC summary doesn’t mention it. |
| LLM -generated | D96BECFB360CDD | Omission | 4 | Did not mention the need to start PJP/GI ppx. |
| LLM -generated | DDBFF0CC359B4E | Inaccuracy | 4 | Discharge summary states inaccurately that patient was receiving chemotherapy while in the hospital (he only received Avastin), saying "The patient's underlying condition of metastatic NSCLC was managed with ongoing chemotherapy and immunotherapy. He was on a regimen of carboplatin, pemetrexed, and pembrolizumab." |
| LLM -generated | D81108FE9360D3 | Omission | 6 | No discussion regarding potential outpatient Surgical Oncology consideration for resection which could be life changing/saving. |
| LLM -generated | D2D74732781DCF | Omission | 6 | DC summary omits concern for pancreatic cancer or T1DM. |
| Physician-generated | D1DF9EF3957287 | Omission | 4 | CT showed aortic ectasia which is not mentioned in the dc summary (also not mentioned in progress notes). |
| Physician-generated | D4641CE4785D03 | Omission | 4 | Omitted transplant candidacy – not currently a candidate due to current active tobacco use. This is important for outpatient providers to know to help encourage tobacco cessation. |
| Physician-generated | D4641CE4785D03 | Omission | 4 | Omitted Smoking Cessation team consult note. Patient was counseled on smoking cessation and this would be helpful to note in the discharge summary. |
| Physician-generated | D67CDEFA8FE96C | Omission | 4 | Patient is noted in encounter notes to have consider EtOH consumption, but this is not discussed in DC summary. If this is not known to PCP, then this may have been a missed opportunity to refer patient to AA or to PCP for further discussion. |
| Physician-generated | D67CDEFA8FE96C | Omission | 4 | Patient is noted in encounter notes to have considerable EtOH consumption and initially elevated transaminases, sufficient that a hepatic US was conducted. Although it appears that transaminitis improves, this is perhaps an important opportunity for follow up by PCP. |

**Supplementary Table 5**. Breakdown of errors assigned a potential harmfulness score (using the adapted AHRQ Common Format Harm scale) of 4 or greater (4 – Potential for permanent harm (lifelong bodily or psychological injury or increased susceptibility to disease), 5 – Potential for lifelong bodily or psychological injury or disfigurement, 6 – Potential for severe permanent harm, 7 – Potential for death).

| **Metric** | **Spearman’s coefficient (p value)** | | | |
| --- | --- | --- | --- | --- |
|  | *Comprehensiveness* | *Concision* | *Coherence* | *Overall quality score* |
| *Cosine similarity* | 0.05 (0.628) | -0.06 (0.532) | 0.05 (0.617) | 0.11 (0.260) |
| *ROUGE-L* | 0.11 (0.293) | 0.16 (0.109) | 0.11 (0.282) | 0.11 (0.297) |
| *BLEU* | 0.22 (0.028) | 0.07 (0.493) | -0.07 (0.504) | 0.16 (0.102) |
| *METEOR* | 0.26 (0.008) | 0.01 (0.915) | -0.08 (0.414) | 0.16 (0.118) |

**Supplementary Table 6.** Spearman’s rank coefficient for the correlation between comprehensiveness, concision, coherence and overall quality scores assigned by reviewers for each LLM-generated summary and the cosine similarity, ROUGE-L, BLEU and METEOR scores calculated between the LLM-generated summary and its physician-generated counterpart.

| **Metric** | **Mean (SD) score** | | |
| --- | --- | --- | --- |
|  | *a) LLM-generated summary compared to physician-generated summary for same encounter* | *b) LLM-generated summary compared to physician-generated summary for random other encounter* | *p value** |
| *Cosine similarity* | 0.94 (0.02) | 0.83 (0.03) | < 0.001 |
| *ROUGE-L* | 0.35 (0.05) | 0.23 (0.04) | < 0.001 |
| *BLEU* | 0.09 (0.05) | 0.05 (0.03) | < 0.001 |
| *METEOR* | 0.27 (0.07) | 0.20 (0.06) | < 0.001 |

**Supplementary Table 7.** Mean (SD) cosine similarity, ROUGE-L, BLEU and METEOR scores for LLM- vs physician-generated summary narratives for a) the same encounter, and b) compared to another randomly selected encounter. *Mann-Whitney U test; p<0.05 considered significant.

**Supplementary Figures**

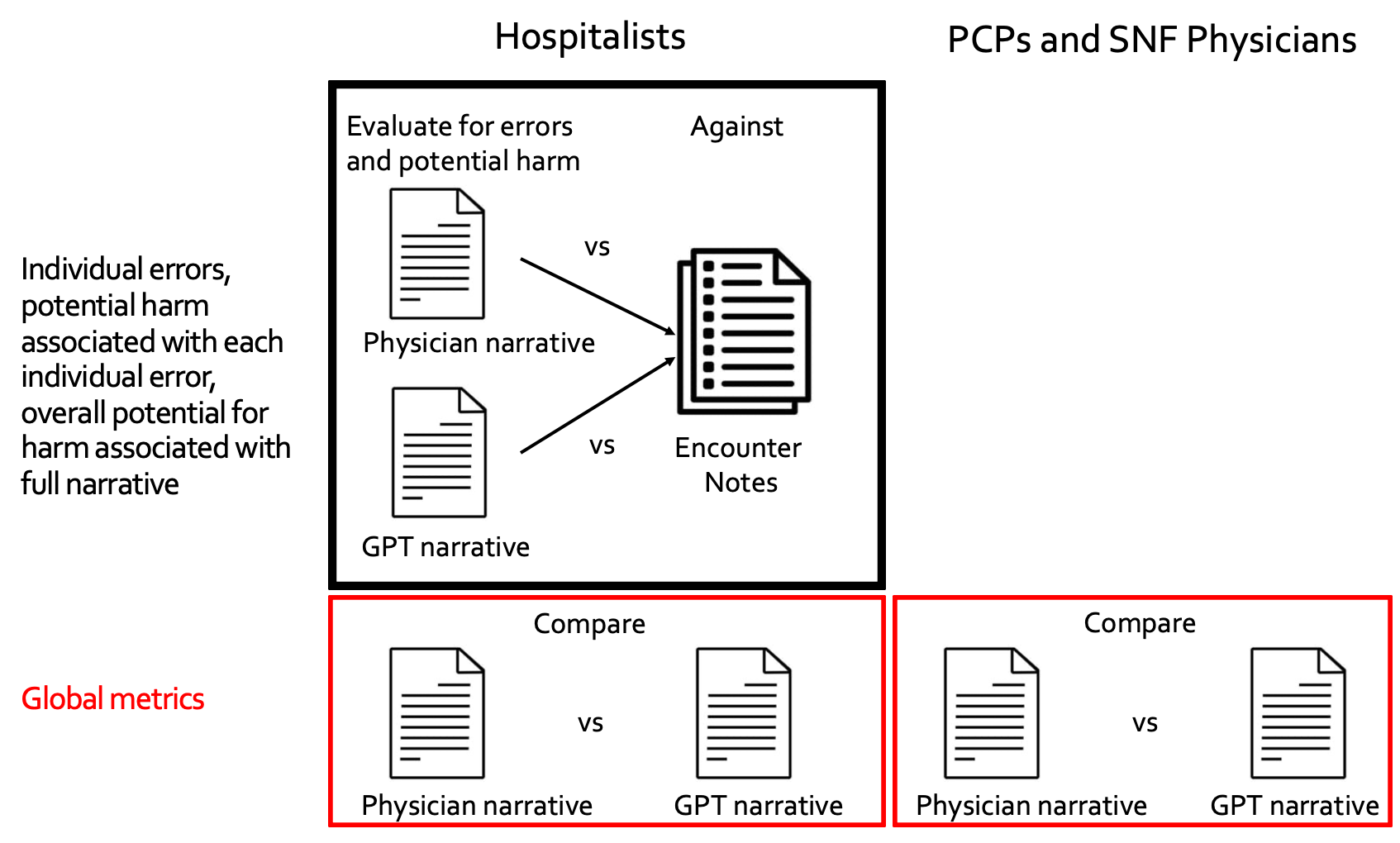
**Supplementary Figure 1.** Metrics evaluated by each reviewer type. PCP = Primary Care Physician; SNF = Skilled Nursing Facility physician.

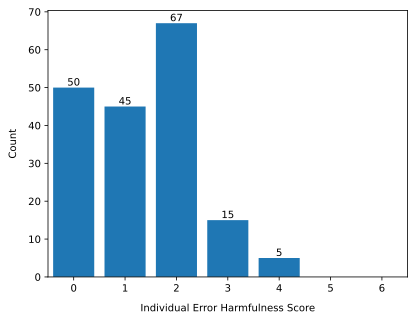

a)

b)

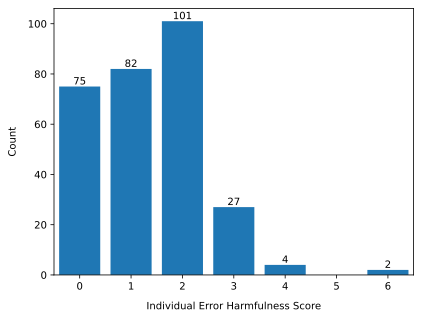

**Supplementary Figure 2**. Histogram of counts of individual error harmfulness scores for a) physician-generated summary narratives and b) LLM-generated summary narratives.

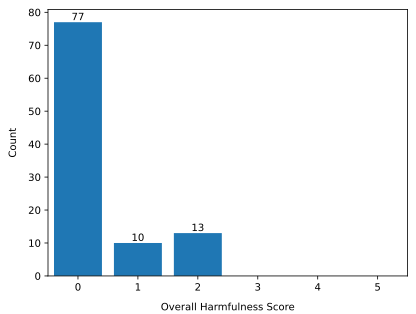

a)

b)

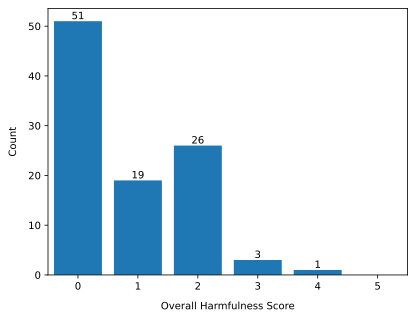

**Supplementary Figure 3**. Histogram of narrative-level, global harmfulness scores for a) physician-generated summary narratives and b) LLM-generated summary narratives.

**Approach to assessing reviewer consistency**

Because hospitalists were tasked with documenting errors by comparing each narrative against the full corpus of hospital encounter notes, we sought to optimize reviewer consistency across three domains: error count, error classification (inaccuracy, omission, and hallucination), and potential for harm from each error.

Before beginning their reviews of the study cohort encounters, all hospitalists underwent training consisting of reviewing three documents from each of two example hospital encounter cases from the development set: 1) the physician-generated narrative, 2) the LLM-generated narrative, and 3) the associated corpus of hospital encounter notes from which narratives were generated. Because error classification inherently involves some degree of subjectivity (e.g. for omission errors, reviewers were instructed to consider whether *in their opinion* an omission would be *material to the PCP or SNF physician’s provision of ongoing care*), there could be no single truth about what constituted all errors. Instead, we identified reviewers whose error counts or individual error potential harmfulness scores were greater than 1 standard deviation from the mean of counts/individual error potential harmfulness scores of all reviewers who had identified the error. We also identified reviewers who misclassified error types in these example cases.

After completing reviews of both narratives (LLM- and physician-generated) for each of the two example cases, reviewers had identified a total of 24 unique errors against the hospital encounter notes. How often a reviewer was ≥ 1 SD from the mean of error counts, harmfulness scores for each error, and whether they had misclassified any error type (e.g. classifying an inaccuracy as an omission) defined whether they were noted to be an outlier on any given occasion (See Protocol Supplementary File 1).

- With respect to error counts, there were 5 reviewers noted as outliers. Three reviewers had error counts ≥1 SD below the mean, while two reviewers had error counts ≥1 SD above the mean (driven mainly by omissions).
- With respect to error type misclassifications, two reviewers were noted as outliers with 1 misclassification error each (inaccuracy error misclassified as omission in both cases).
- With respect to potential harmfulness scores, four reviewers were noted as outliers with individual error harmfulness scores ≥ 1 SD from the mean.

Outlier reviewers were re-trained as specified in the Study Protocol.

Following completion of reviewer training, each of the 100 study cohort encounters were reviewed by two independent hospitalist reviewers, resulting in 400 narrative reviews for errors and harmfulness in total (100 LLM- and 100 physician-generated narratives each reviewed in duplicate). Duplicate errors were consolidated by a 3^rd^ party adjudicator, who also removed any errors which met error exclusion criteria (e.g. items mistakenly reported as omissions from the narrative which normally fall under non-narrative headings elsewhere in the full discharge summary, such as pending labs; see Supplementary File 1 for further details) and re-classified any misclassified error types. Following consolidation, 473 unique errors were identified across all reviews, with 10 errors misclassified, giving a 2.1% error misclassification rate.
